# Conversational Artificial Intelligence-Enabled Molecular Characterization of Sézary Syndrome Reveals Distinct Pathway-Level Alterations Compared with Non-Sézary Cutaneous T-Cell Lymphoma

**DOI:** 10.64898/2026.03.09.26347970

**Authors:** Fernando C. Diaz, Brigette Waldrup, Francisco G. Carranza, Sophia Manjarrez, Enrique Velazquez-Villarreal

## Abstract

**Background:** Sézary syndrome (SS) represents an aggressive leukemic variant of cutaneous T-cell lymphoma (CTCL) with distinct clinical behavior compared with other CTCL subtypes. While prior studies have identified recurrent genomic alterations in CTCL, a systematic pathway-centric comparison between SS and non-SS CTCL remains limited. We applied our conversational artificial intelligence (AI) platform for precision oncology, to accelerate hypothesis generation and integrative interpretation of public genomic data.

**Methods:** We performed a secondary analysis of somatic mutation and clinical data from the Columbia University CTCL cohort available via cBioPortal. Samples were stratified into SS (n=26) and non-SS CTCL (n=17). High-impact coding variants were retained and annotated to curated functional gene groups and signaling pathways relevant to CTCL biology. Pathway-level mutation frequencies were compared using Fisher’s exact test, with effect sizes summarized by odds ratios. Tumor mutation burden (TMB) was compared using Wilcoxon rank-sum testing. Subtype-specific gene-gene co-mutation patterns were assessed using pairwise association testing and visualized with heatmaps and oncoplots, with our conversational AI agents facilitating interactive exploration and prioritization of results.

**Results:** Overall TMB did not differ between SS and non-SS CTCL (p=0.83), indicating comparable global mutational burden. Pathway-level analyses revealed enrichment of alterations affecting epigenetic regulators, tumor suppressor and cell-cycle control genes, NFAT signaling, and apoptosis/immune regulation in SS, whereas MAPK and JAK-STAT pathway alterations were relatively more frequent in non-SS CTCL. Co-mutation analysis demonstrated fewer but more focused gene-gene interactions in SS compared with broader co-mutation networks in non-SS CTCL, suggesting divergent evolutionary constraints. Several genes (including ERBB2, WWC1, POSTN) showed borderline subtype-specific enrichment, warranting further validation.

**Conclusions:** Conversational AI-enhanced analysis reveals that SS is distinguished from other CTCL subtypes not by higher mutational load, but by qualitative differences in pathway involvement, particularly epigenetic dysregulation, immune escape, and transcriptional control. These findings generate testable hypotheses for downstream validation in patient-level datasets and demonstrate the utility of conversational AI agents as accelerators of translational cancer genomics.

## 1. Introduction

Cutaneous T-cell lymphoma (CTCL) comprises a heterogeneous group of extranodal non-Hodgkin lymphomas characterized by the clonal expansion of malignant skin-homing CD4⁺ T lymphocytes. Although CTCL is a rare malignancy, it represents a clinically significant disease due to its chronic course, complex diagnosis, and potential progression to advanced systemic involvement. Epidemiologic analyses based on population-level datasets indicate that more than 14,000 individuals were diagnosed with CTCL in the United States between 2000 and 2018, with incidence trends showing gradual increases over recent decades (1). Overall, CTCL accounts for approximately 4% of non-Hodgkin lymphomas and about 0.14% of all cancers, underscoring its rarity while highlighting the need for deeper molecular characterization and improved therapeutic strategies (2–6). Despite advances in genomic profiling and targeted therapies, CTCL remains a biologically complex disease with substantial clinical heterogeneity (7–9).

CTCL encompasses multiple clinicopathologic subtypes, among which mycosis fungoides (MF) and Sézary syndrome (SS) represent the most widely recognized entities. MF is the most common subtype and typically follows a relatively indolent course characterized by slowly evolving skin lesions that may progress from patch and plaque stages to tumor formation or systemic disease in a subset of patients (7). In contrast, SS represents an aggressive leukemic variant defined by erythroderma, lymphadenopathy, and the presence of circulating malignant T cells, and is associated with markedly poorer clinical outcomes (7). Despite belonging to the same disease spectrum, MF and SS display substantial differences in clinical presentation, disease progression, and therapeutic response, suggesting that distinct molecular programs underlie these CTCL subtypes. Early-stage disease can be particularly challenging to diagnose because CTCL often mimics benign inflammatory dermatologic conditions such as eczema, psoriasis, or chronic dermatitis, frequently delaying definitive diagnosis and treatment (8,10).

Management of CTCL remains complex and requires a multidisciplinary approach involving dermatology, hematology-oncology, pathology, and radiation oncology. Treatment strategies vary widely depending on disease stage and subtype and may include skin-directed therapies, systemic immunomodulatory agents, targeted therapies, and radiation therapy (7,8,10,11). Although several therapeutic options have improved patient outcomes, durable curative therapies remain limited, and many patients experience disease relapse or progression over time (7). Emerging treatments such as immune checkpoint blockade and targeted biologics, including anti-PD-L1-based immunotherapy or monoclonal antibodies such as mogamulizumab, have shown promise in selected CTCL populations but highlight the need for improved molecular stratification to guide precision treatment approaches (12,13).

The molecular pathogenesis of CTCL is incompletely understood and is thought to involve a combination of genomic alterations, dysregulated immune signaling, and microenvironmental interactions. Genomic studies have identified recurrent somatic mutations affecting signaling pathways involved in T-cell receptor signaling, MAPK signaling, RAS pathway activation, epigenetic regulation, and cytokine-mediated pathways such as JAK-STAT (14–19). Integrated genomic analyses of CTCL cohorts have revealed substantial molecular heterogeneity, suggesting that distinct genetic programs may drive different clinical phenotypes across CTCL subtypes (16,20). More recent single-cell and spatial analyses have further highlighted the complex tumor ecosystem of CTCL, revealing malignant TH2-skewed T-cell populations interacting with B-cell-rich tumor microenvironments and immune regulatory networks that contribute to disease progression (21–23). These findings underscore the importance of studying CTCL biology at both the genomic and microenvironmental levels.

Despite these advances, most genomic studies of CTCL have focused primarily on individual gene mutations or subtype-specific analyses, and systematic pathway-level comparisons between SS and other CTCL subtypes remain limited. Because oncogenic processes often arise through coordinated dysregulation of signaling networks rather than isolated gene mutations, pathway-centered analyses may provide a more biologically informative framework for understanding subtype-specific disease mechanisms and identifying therapeutic vulnerabilities.

Recent developments in artificial intelligence (AI) and computational oncology have created new opportunities to accelerate discovery in translational cancer genomics. AI-driven analytical frameworks can facilitate exploration of large-scale genomic datasets, enable interactive interrogation of complex biological relationships, and support hypothesis generation in precision medicine research (24,25). Among these approaches, conversational AI systems have emerged as novel tools capable of integrating genomic, clinical, and biological knowledge to guide exploratory data analysis and interpret multidimensional datasets.

In this study, we applied AI-HOPE (26), a conversational AI framework designed to integrate genomic and clinical datasets for precision oncology research, AI-HOPE-JAK-STAT (27) and AI-HOPE-MAPK (28). AI-HOPE enables dynamic cohort interrogation, pathway-level analysis, and interactive hypothesis generation through natural language-guided analytical workflows. By facilitating rapid exploration of complex genomic repositories such as cBioPortal, AI-HOPE supports pathway-centric interpretation of mutational landscapes while complementing conventional statistical analyses.

Using this AI-enabled analytical approach (29,30), we performed a secondary analysis of genomic and clinical data from the Columbia University CTCL cohort to systematically compare the somatic mutational landscape of SS and non-SS CTCL (3,31–35). Through pathway-level aggregation of somatic mutations and evaluation of gene-gene interaction patterns, this study aims to identify subtype-specific molecular signatures and signaling dependencies that distinguish SS from other CTCL variants. By integrating pathway-centric genomic analysis with conversational AI-assisted exploration, our work seeks to improve understanding of CTCL molecular heterogeneity and generate new hypotheses for precision oncology strategies in this rare but clinically challenging malignancy.

## 2. Methods

### 2.1 Data Source and Cohort Definition

This study represents a secondary analysis designed to molecularly characterize and compare the somatic mutational landscape of SS and non-SS CTCL using publicly available genomic and clinical datasets. Mutation and clinical annotation data were obtained from the Columbia University CTCL cohort accessible through the cBioPortal for Cancer Genomics platform. cBioPortal provides harmonized genomic datasets derived from multiple institutional studies and allows standardized analysis of somatic mutation and clinical features across cancer cohorts.

Tumor samples were stratified into two groups based on the detailed cancer type annotation within the dataset. The SS consisted of samples annotated specifically as SS (n = 26). The non-SS CTCL cohort comprised all remaining CTCL subtypes (n = 17), including MF (n = 3), primary cutaneous CD30-positive T-cell lymphoproliferative disorders (n = 1), and primary cutaneous CD8-positive aggressive epidermotropic cytotoxic T-cell lymphoma (n = 13). All samples were classified within the broader disease category of mature T– and NK-cell neoplasms. This stratification enabled direct comparison of genomic alterations between the leukemic CTCL subtype SS and other CTCL variants.

### 2.2 Mutation Data Processing and Filtering

Somatic mutation data were retrieved directly from the cBioPortal mutation annotation files associated with the Columbia CTCL dataset. To focus analyses on biologically meaningful alterations with potential functional consequences, only high-impact coding variants were retained for downstream analyses. High-impact variants were defined according to variant classification categories commonly associated with functional protein changes. These included: Missense mutations, Nonsense mutations, Frameshift insertions and deletions, Splice-site mutations, In-frame insertions and deletions, and Translation start-site alterations

This filtering strategy enriched the dataset for variants most likely to influence protein structure, gene function, and downstream signaling pathways relevant to CTCL biology.

### 2.3 Gene-Level and Pathway-Level Mutation Annotation

Filtered somatic variants were annotated to predefined functional gene groups and signaling pathways based on established biological roles in CTCL pathogenesis and T-cell malignancies. Gene group assignments were curated from previously reported CTCL genomic studies (3) and canonical pathway annotations derived from established biological databases (3).

The following functional gene groups and signaling pathways were analyzed: epigenetic regulators (TET2, DNMT3A, CREBBP, KMT2D, KMT2C, ARID1A, SMARCA4, CHD3, and BRD9), which are involved in chromatin remodeling, DNA methylation, and transcriptional regulation; tumor suppressor genes (TP53, RB1, PTEN, CDKN1B, and CDKN2A), critical for maintaining genomic stability, regulating cell-cycle checkpoints, and controlling apoptotic signaling; cell-cycle regulators (TP53, RB1, CDKN1B, and CDKN2A), which govern key checkpoints in cell-cycle progression; T-cell receptor (TCR) signaling components (PLCG1, VAV1, LAT, LCK, ZAP70, and FYN), mediating proximal TCR activation and downstream signaling cascades; the JAK–STAT signaling pathway (JAK1, JAK3, STAT3, STAT5B, and SOCS1), which regulates cytokine-mediated signaling and immune activation; the MAPK signaling pathway (MAPK1, MAPK3, BRAF, KRAS, NRAS, RAF1, MAP2K1, MAP2K2, NF1, and RASA1), representing central nodes of the mitogen-activated protein kinase cascade; the NF-κB signaling pathway (CARD11, TNFAIP3, IKBKB, CHUK, NFKB1, NFKB2, REL, RELA, and RELB), which controls inflammatory signaling and T-cell survival; the NFAT signaling pathway (NFATC1, NFATC2, PRKG1, PPP3CA, PPP3CB, and PPP3R1), involved in calcium-dependent transcriptional activation and immune signaling; DNA damage response genes associated with genomic integrity and DNA repair processes, including TP53 and related components; and genes involved in apoptosis and immune regulation (FAS, FASLG, BCL2, BCL6, PDCD1, and CTLA4), which regulate programmed cell death, immune checkpoint signaling, and tumor immune evasion.

For each tumor sample, a functional gene group or signaling pathway was classified as mutated if at least one high-impact somatic variant was present in any gene belonging to that pathway. Pathway mutation status was therefore represented as a binary variable (mutated vs. wild type) for downstream statistical comparisons between cohorts.

### 2.4 Pathway-Level Statistical Analysis

Mutation frequencies for each functional gene group and signaling pathway were compared between the SS and non-SS CTCL cohorts. Statistical comparisons were performed using Fisher’s exact test, which is appropriate for categorical comparisons in small sample sizes.

To quantify the magnitude and direction of associations between pathway alterations and CTCL subtype, odds ratios (ORs) and corresponding confidence intervals were estimated. Results were visualized using forest plots to facilitate interpretation of pathway-level enrichment patterns across CTCL subtypes.

Gene-level mutation frequencies were also calculated for each cohort, and Fisher’s exact test was used to identify genes with potential subtype-specific enrichment. Genes demonstrating borderline statistical significance were highlighted for exploratory interpretation and hypothesis generation.

### 2.5 Tumor Mutation Burden Analysis

Tumor mutation burden (TMB) values were obtained directly from the clinical annotation files provided in the cBioPortal dataset. TMB represents the total number of somatic mutations per tumor sample and serves as a surrogate measure of global mutational load.

TMB distributions between SS and non-SS CTCL cohorts were compared using boxplot visualization and statistical testing with the Wilcoxon rank-sum test. This nonparametric test was selected because it does not assume normal distribution of mutation counts and is appropriate for small cohort sizes.

### 2.6 Co-Mutation Analysis

To evaluate subtype-specific patterns of mutational interaction, pairwise gene-gene co-mutation analyses were performed using the most frequently mutated genes in the dataset. Analyses were conducted separately for the SS and non-SS cohorts to identify patterns of mutational co-occurrence or mutual exclusivity.

Statistical testing of gene-gene associations was performed using Fisher’s exact test. Results were visualized as co-mutation heatmaps, enabling identification of recurrent interaction patterns among mutated genes within each CTCL subtype.

### 2.7 Visualization of Mutational Landscape

Multiple visualization approaches were used to summarize the genomic landscape of CTCL subtypes: Oncoplots were generated to display somatic mutation patterns across the most frequently altered genes in the cohort. Each column represents an individual tumor sample and each row represents a gene. Colored blocks indicate the presence and classification of high-impact mutations. Lollipop plots were generated for the top recurrently mutated genes in each cohort to illustrate mutation positions along the protein sequence and identify potential mutation hotspots. Forest plots were used to summarize pathway-level mutation enrichment and associated odds ratios between cohorts.

### 2.8 Conversational Artificial Intelligence-Assisted Analysis

A conversational artificial intelligence (AI) platform developed for precision oncology research was used to facilitate interactive exploration, prioritization, and interpretation of genomic findings. The AI framework enabled rapid querying of genomic datasets, dynamic visualization of mutation patterns, and generation of pathway-level hypotheses by integrating mutation data, biological pathway annotations, and statistical outputs.

This AI-assisted workflow served as a complementary analytical layer to conventional statistical methods, accelerating hypothesis generation and enabling efficient interpretation of complex genomic data while maintaining transparency and reproducibility of analytical steps.

## 3. Results

### 3.1 Cohort composition and clinicopathologic stratification

The study cohort comprised 43 CTCL tumors from the Columbia University dataset available through cBioPortal, including 26 SS cases and 17 non-SS CTCL cases (Table 1). All samples were classified within the broader category of mature T– and NK-cell neoplasms. The non-SS group was heterogeneous and included MF (n = 3), primary cutaneous CD30-positive T-cell lymphoproliferative disorders (n = 1), and predominantly primary cutaneous CD8-positive aggressive epidermotropic cytotoxic T-cell lymphoma (n = 13). This framework enabled direct comparison between the leukemic SS phenotype and other clinically distinct CTCL subtypes.

**Table 1.** Clinical and histopathologic characteristics of the Sézary syndrome and non-Sézary cutaneous T-cell lymphoma cohorts analyzed in the Columbia University CTCL dataset.

| Clinical Feature | Sézary Syndrome Cohort<br>n (%) | Non-Sézary Syndrome Cohort<br>n (%) |
| --- | --- | --- |
| **Cancer Type** |  |  |
| Mature T and NK Neoplasms | 26<br>(100.0%) | 17<br>(100.0%) |
| **Detailed Cancer Type** |  |  |
| Mycosis Fungoides | 0 (0.0%) | 3 (17.6%) |
| Primary Cutaneous CD30 Positive T-Cell Lymphoproliferative Disorders | 0 (0.0%) | 1 (5.9%) |
| Primary Cutaneous CD8 Positive Aggressive Epidermotropic Cytotoxic T-Cell Lymphoma | 0 (0.0%) | 13<br>(76.5%) |
| Sézary Syndrome | 26<br>(100.0%) | 0 (0.0%) |

### 3.2 Comparable tumor mutational burden in Sézary syndrome and non-Sézary CTCL

We first examined whether SS was distinguished from non-SS CTCL by differences in overall mutational burden (Fig. 1). TMB obtained from the cBioPortal clinical annotations, showed substantial inter-sample variability in both groups, but the overall distributions were highly comparable. Median TMB values and interquartile ranges overlapped extensively between SS and non-SS CTCL, and Wilcoxon rank-sum testing demonstrated no significant difference between cohorts (P = 0.83). These data indicate that the molecular distinction between SS and non-SS CTCL is not driven by a higher global mutational load in SS, but instead is more likely attributable to qualitative differences in the identity and pathway context of somatic alterations.

**Figure 1.**
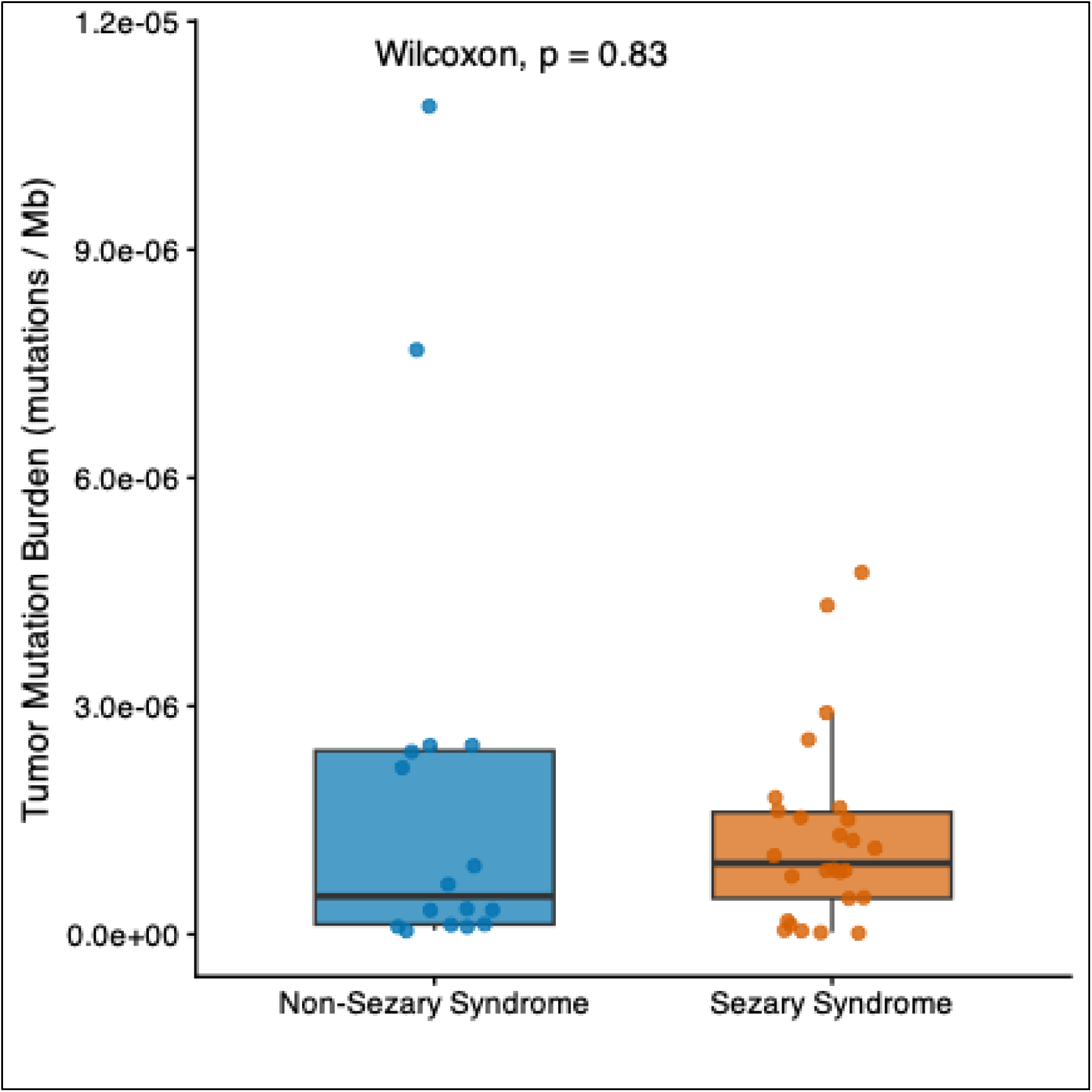
Comparable tumor mutational burden between Sézary syndrome and non-Sézary cutaneous T-cell lymphoma.

### 3.3 Sézary syndrome exhibits preferential involvement of epigenetic, tumor suppressor and NFAT-associated programs

To determine whether specific biological pathways were differentially altered across CTCL subtypes, we aggregated high-impact coding variants into predefined functional gene groups and signaling modules relevant to T-cell lymphoma biology (Table 2). Pathway-level comparisons revealed a consistent pattern in which SS showed greater involvement of pathways linked to epigenetic regulation, tumor suppressor dysfunction, cell-cycle control, NFAT signaling, and apoptosis/immune regulation, whereas MAPK and JAK-STAT pathway alterations were relatively more frequent in non-SS CTCL.

**Table 2.** Pathway-level comparison of somatic mutation frequencies between Sézary syndrome and non-Sézary cutaneous T-cell lymphoma cohorts.

| Functional Group or Pathway | Sézary Syndrome Mutated n (%) | Sézary Syndrome Wild-Type n (%) | Non-Sézary Syndrome Mutated n (%) | Non-Sézary Syndrome Wild-Type n (%) | p-value |
| --- | --- | --- | --- | --- | --- |
| Epigenetic Regulation | 10 (38.5%) | 16 (61.5%) | 3 (17.6%) | 14 (82.4%) | 0.1874 |
| Tumor Suppressor Pathway | 4 (15.4%) | 22 (84.6%) | 1 (5.9%) | 16 (94.1%) | 0.6327 |
| Cell Cycle Regulation | 4 (15.4%) | 22 (84.6%) | 1 (5.9%) | 16 (94.1%) | 0.6327 |
| T-Cell Receptor Signaling | 3 (11.5%) | 23 (88.5%) | 1 (5.9%) | 16 (94.1%) | 1 |
| JAK/STAT Signaling | 1 (3.8%) | 25 (96.2%) | 2 (11.8%) | 15 (88.2%) | 0.5523 |
| MAPK Signaling | 2 (7.7%) | 24 (92.3%) | 3 (17.6%) | 14 (82.4%) | 0.3686 |
| NF-κB Signaling | 2 (7.7%) | 24 (92.3%) | 1 (5.9%) | 16 (94.1%) | 1 |
| NFAT Signaling | 4 (15.4%) | 22 (84.6%) | 1 (5.9%) | 16 (94.1%) | 0.6327 |
| DNA Damage Response | 4 (15.4%) | 22 (84.6%) | 2 (11.8%) | 15 (88.2%) | 1 |
| Apoptosis and Immune Regulation | 1 (3.8%) | 25 (96.2%) | 0 (0.0%) | 17 (100.0%) | 1 |

Among the pathway groups enriched in SS, epigenetic regulators were altered in 10 of 26 SS tumors (38.5%) compared with 3 of 17 non-SS tumors (17.6%). Similarly, tumor suppressor pathway alterations were present in 15.4% of SS cases versus 5.9% of non-SS cases, and the same distribution was observed for the overlapping cell-cycle regulation category. NFAT pathway alterations were also more frequent in SS (15.4% versus 5.9%), as were T-cell receptor signaling alterations (11.5% versus 5.9%). Notably, alterations in apoptosis and immune regulation were observed only in SS (3.8% versus 0%), yielding the strongest directional effect favoring the SS subtype despite the low absolute event rate.

In contrast, MAPK pathway mutations were more frequent in non-SS CTCL (17.6%) than in SS (7.7%), and JAK-STAT pathway alterations followed a similar pattern (11.8% versus 3.8%). NF-κB signaling and DNA damage response genes showed relatively similar frequencies across subtypes, suggesting that these pathways may represent shared molecular features of CTCL rather than subtype-defining programs.

Although most pathway-level comparisons did not reach nominal statistical significance, likely reflecting the modest cohort size, effect-size analyses using odds ratios demonstrated clear directional divergence between the two disease groups. Forest plot visualization showed odds ratios greater than one for epigenetic regulation, tumor suppressor genes, cell-cycle control, NFAT signaling, and apoptosis/immune regulation, consistent with enrichment in SS, whereas odds ratios below one were observed for MAPK and JAK-STAT signaling, indicating relative enrichment in non-SS CTCL (Figure S1). Collectively, these data support a model in which SS is characterized by preferential perturbation of transcriptional and immune-regulatory circuitry rather than by increased mutational burden per se.

### 3.4 Gene-level analysis identifies recurrent subtype-associated alterations and candidate discriminatory genes

At single-gene resolution, recurrent alterations were distributed differently across the two CTCL groups (Table 3). In the SS cohort, recurrently mutated genes included TET2, PLCG1, and TP53, each altered in 3 of 26 tumors (11.5%), as well as PRKG1, CREBBP, CHD3, and CARD11, each mutated in 2 tumors (7.7%). Additional SS-associated events included mutations in STAT5B, BRD9, NF1, MAPK3, and PDCD1. This pattern reinforced the pathway-level observation that SS is enriched for alterations involving epigenetic machinery, T-cell signaling, transcriptional control, tumor suppressor loss and immune-regulatory genes.

**Table 3.**
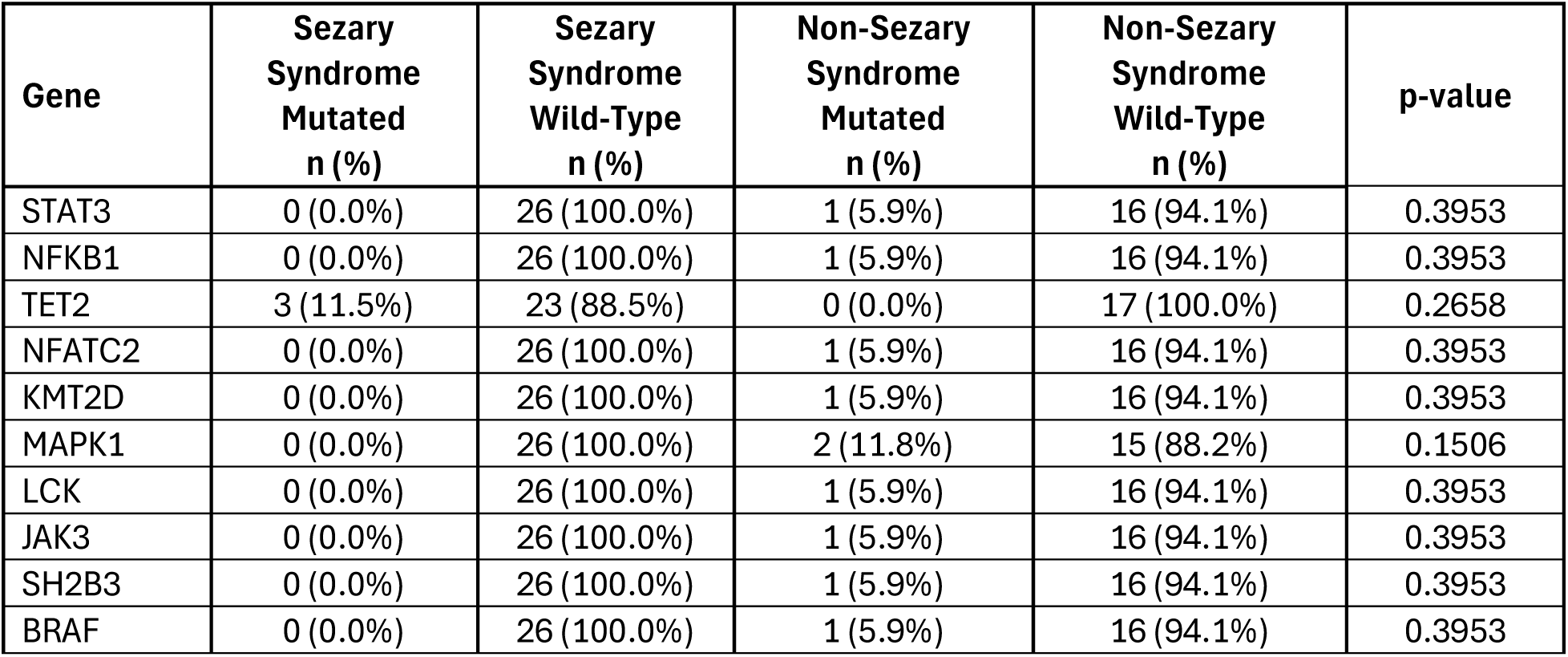
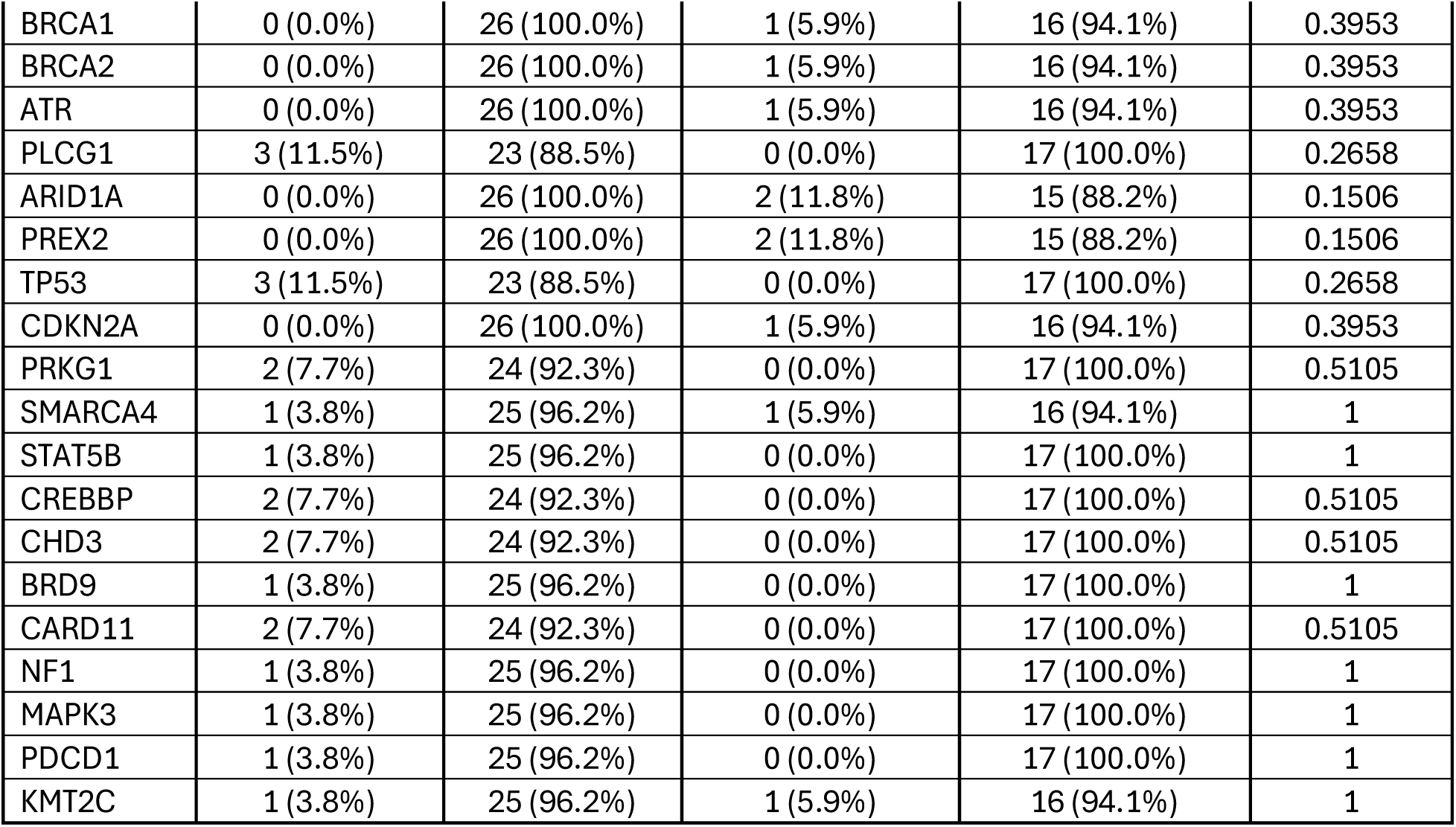
Gene-level comparison of somatic mutation frequencies between Sézary syndrome and non-Sézary cutaneous T-cell lymphoma cohorts.

By contrast, the non-SS cohort showed recurrent mutations in genes associated with signaling diversity and broader pathway dispersion. MAPK1, ARID1A, and PREX2 were each mutated in 2 of 17 tumors (11.8%), while single events were observed in STAT3, NFKB1, NFATC2, KMT2D, LCK, JAK3, SH2B3, BRAF, BRCA1, BRCA2, ATR, and CDKN2A. These findings suggest that non-SS CTCL harbors a wider distribution of low-frequency mutations affecting MAPK, JAK-STAT, NF-κB, DNA damage and chromatin-associated genes, rather than the more concentrated recurrent pattern seen in SS.

We next examined genes showing borderline subtype-specific enrichment (Table S1). Several genes, including ERBB2, GUCY2F, WWC1, PCDHB13, and POSTN, were mutated in 3 of 17 non-SS tumors (17.6%) but in none of the 26 SS tumors, each yielding P = 0.0551 by Fisher’s exact test. Although these associations did not cross conventional significance thresholds, their consistent enrichment in non-SS CTCL suggests that they may represent biologically relevant subtype-associated candidates that warrant validation in larger cohorts. Particularly notable among these were ERBB2, a canonical receptor tyrosine kinase with established oncogenic activity in multiple tumor types, and POSTN, which has been implicated in extracellular matrix remodeling and tumor-stromal interactions.

### 3.5 Distinct co-mutation architecture differentiates SS from non-SS CTCL

Because pathway-level differences may reflect not only the presence of mutations but also their combinatorial organization, we evaluated pairwise gene-gene co-mutation patterns separately in SS and non-SS CTCL (Fig. 2). This analysis revealed marked differences in network architecture between subtypes.

**Figure 2.**
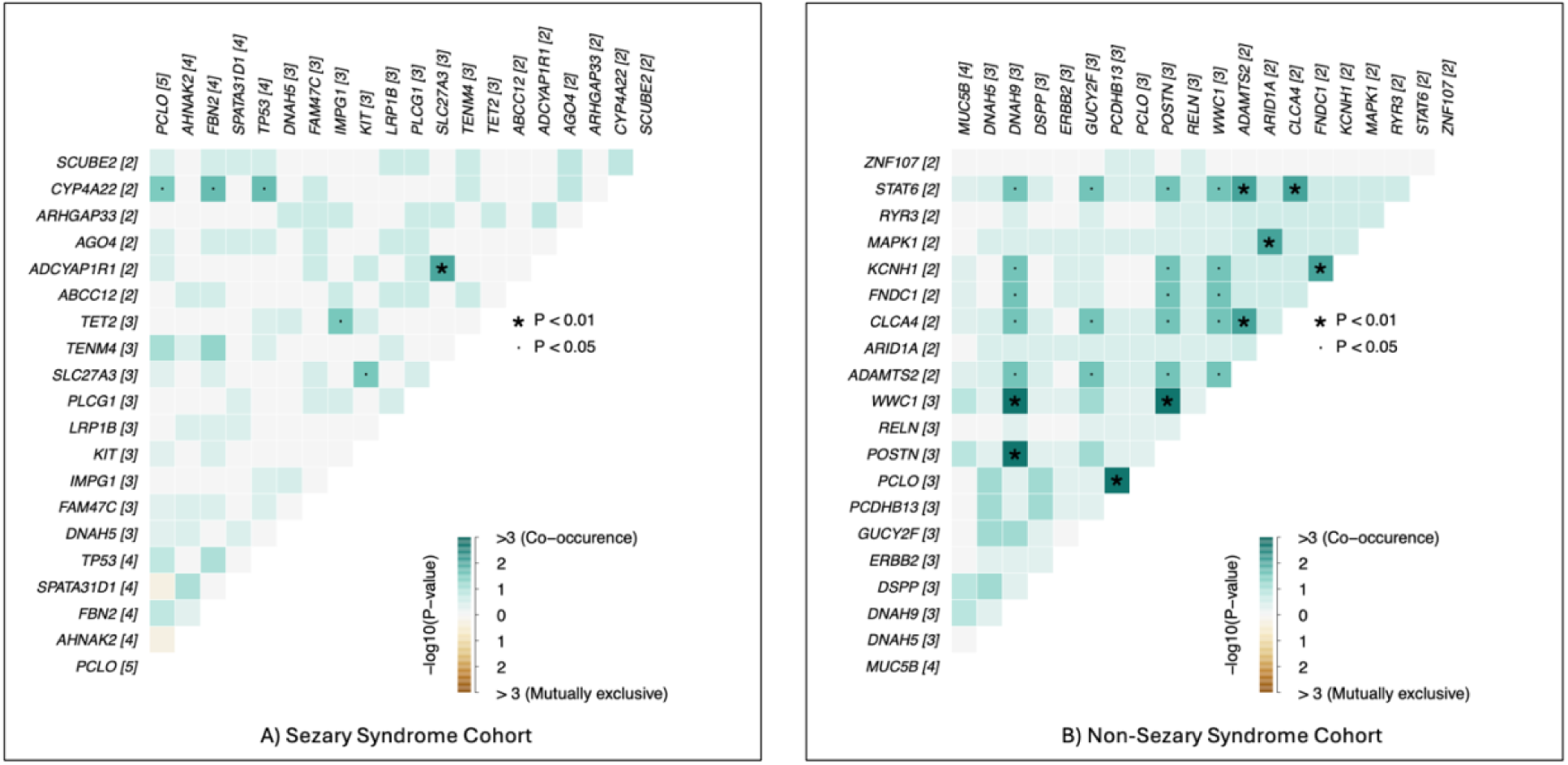
Distinct gene-gene mutational interaction patterns in Sézary syndrome and non-Sézary cutaneous T-cell lymphoma. Subtype-stratified gene-gene co-mutation heatmaps were constructed for the 20 most frequently mutated genes in Sézary syndrome (SS) and non-Sézary (non-SS) cutaneous T-cell lymphoma (CTCL) cohorts. Pairwise mutational interactions were evaluated using Fisher’s exact test to identify statistically significant co-occurrence or mutual exclusivity relationships across tumor samples. The SS cohort (panel A) demonstrated a relatively limited interaction network, with six significant gene-gene associations detected. In contrast, the non-Sézary (non-SS) CTCL cohort (panel B) exhibited a substantially larger set of significant interactions (27 gene pairs), indicating a broader mutational co-occurrence landscape. Heatmap color intensity reflects the −log10(p-value) of pairwise associations, with positive values representing co-occurrence and negative values indicating mutual exclusivity; asterisks denote statistically significant interactions (P < 0.01), and dots indicate nominal significance (P < 0.05). These results highlight subtype-specific mutational architectures in CTCL, with non-SS tumors displaying more complex gene-gene interaction patterns compared with the relatively constrained mutational network observed in SS.

The SS cohort demonstrated a comparatively restricted co-mutation structure, with only six statistically significant gene-pair interactions identified among the top recurrently mutated genes. These interactions were more focused, suggesting a more constrained mutational architecture in which a smaller set of cooperating lesions may underlie the SS phenotype. This pattern is consistent with the notion that SS may depend on a narrower range of transcriptional, immune and epigenetic dependencies.

In contrast, the non-SS CTCL cohort exhibited a substantially broader co-mutation network, with 27 significant gene-pair interactions detected. The increased number of co-occurring mutational relationships in non-SS CTCL suggests greater genomic heterogeneity and a more distributed architecture of cooperating somatic events. Rather than converging on a limited number of central programs, non-SS CTCL appears to involve a broader range of interacting molecular lesions spanning multiple pathways.

This divergence in co-mutation topology implies different evolutionary constraints across CTCL subtypes. SS may evolve through a more selective and coherent mutational program centered on epigenetic dysregulation, transcriptional control and immune escape, whereas non-SS CTCL may be shaped by a wider mutational repertoire with more diffuse signaling interdependencies.

### 3.6 Oncoplot analysis highlights subtype-specific mutational organization rather than differences in mutation quantity

To visualize the overall mutational landscape at sample-level resolution, we generated an oncoplot of the top 20 recurrently altered genes across the full cohort, ordered by subtype (Fig. 3). This representation confirmed that both SS and non-SS CTCL contained tumors with variable mutational burdens, consistent with the TMB analysis, but also highlighted clear differences in the composition and organization of recurrent alterations.

**Figure 3.**
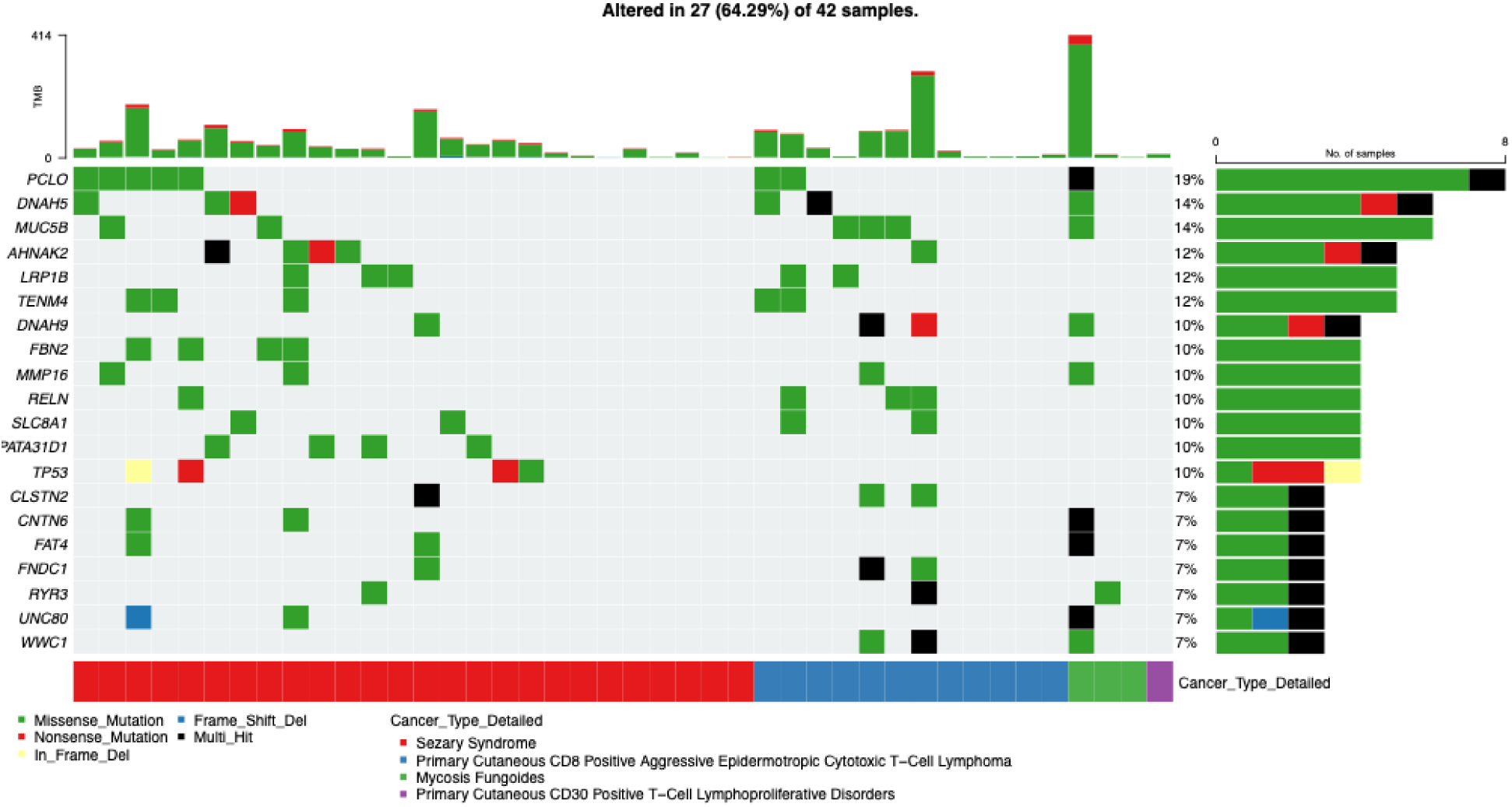
Oncoplot depicting the mutational landscape of Sézary syndrome and non-Sézary cutaneous T-cell lymphoma. This oncoplot summarizes somatic mutation patterns across the 20 most frequently mutated genes identified in the cutaneous T-cell lymphoma (CTCL) cohort. Each column represents an individual tumor sample and each row corresponds to a gene, while colored tiles indicate the presence and type of high-impact somatic alterations detected in the genomic dataset. Samples are arranged by subtype, with Sézary syndrome (SS) tumors displayed on the left and non-Sézary syndrome (non-SS) CTCL tumors on the right. The bar plot above the heatmap represents the tumor mutation burden (TMB) for each sample based on clinical annotations from the dataset. The annotation track below the heatmap indicates the detailed CTCL subtype classification for each tumor. Overall, the oncoplot highlights subtype-specific mutation patterns and heterogeneity across the cohort, illustrating differences in the distribution of recurrent gene alterations and overall mutational profiles between SS and non-SS CTCL.

SS tumors were characterized by recurrent mutations in genes linked to epigenetic regulation (TET2, CREBBP, CHD3, BRD9), tumor suppression and cell-cycle control (TP53), T-cell signaling (PLCG1), and immune regulation (PDCD1). In contrast, non-SS CTCL samples displayed recurrent alterations distributed across MAPK signaling (MAPK1, BRAF), JAK-STAT signaling (STAT3, JAK3), chromatin remodeling (ARID1A, KMT2D) and candidate subtype-associated genes such as ERBB2 and POSTN.

Lollipop plots of the top mutated genes further suggested that mutations were dispersed across protein coding regions rather than clustering into a small number of highly recurrent hotspots, consistent with the heterogeneous and pathway-oriented nature of CTCL mutagenesis (Figs. S2 and S3). Together, these visual analyses reinforced the conclusion that SS and non-SS CTCL are distinguished less by total mutational burden than by the identity, pathway context and combinatorial structure of their somatic alterations.

### 3.7 Conversational AI facilitates prioritization of subtype-associated molecular patterns

Throughout the analytical workflow, the conversational AI framework enabled rapid interrogation of the genomic dataset and facilitated identification of candidate subtype-associated genes and pathway-level alterations across CTCL cohorts. Rather than replacing conventional statistical analysis, the AI-assisted system functioned as an exploratory interface that accelerated hypothesis generation, cohort definition, and visualization of mutation frequency patterns prior to formal statistical evaluation.

To illustrate this capability, we conducted targeted conversational AI-guided case-control analyses evaluating mutation prevalence across selected genes and pathways. For example, ERBB2 mutation frequency was examined by comparing non-SS CTCL tumors (case cohort, n = 17) with SS syndrome samples (control cohort, n = 25). This analysis revealed ERBB2 mutations exclusively in the non-SS CTCL group (3/17 cases) and none in the SS cohort (0/25), suggesting a potential subtype-associated enrichment pattern, although statistical confidence was limited by sample size (Fig. S4).

A similar conversational AI-guided comparison was performed for PCLO mutations, which demonstrated comparable mutation frequencies between cohorts. PCLO alterations were detected in 5 of 25 SS tumors (20.0%) and 3 of 17 non-SS CTCL tumors (17.65%), with Fisher’s exact testing indicating no statistically significant difference between the groups (Fig. S5).

In addition to individual gene comparisons, the AI platform facilitated pathway-level exploration of mutation patterns. Analysis of epigenetic regulatory genes demonstrated a higher frequency of alterations in the SS cohort (10/25; 40.0%) relative to non-SS CTCL (3/17; 17.65%), consistent with the broader pathway enrichment patterns observed in the primary analyses. Although the difference did not reach statistical significance, the calculated odds ratio suggested a directional trend toward epigenetic pathway enrichment in SS (Fig. S6).

Collectively, these conversational AI-guided exploratory analyses enabled rapid construction of case-control comparisons and visualization of mutation distributions across CTCL subtypes. When integrated with standard statistical testing and pathway-based analyses, this approach helped prioritize biologically coherent signals within the dataset. Across the study, this combined framework consistently highlighted a molecular distinction between CTCL subtypes, in which SS exhibited greater involvement of epigenetic regulatory pathways, tumor suppressor programs, and NFAT-linked signaling networks, whereas non-SS CTCL showed relatively greater representation of MAPK and JAK-STAT pathway alterations and broader co-mutation networks.

Taken together, these findings support a model in which SS is characterized by qualitative pathway-level remodeling rather than increased mutational burden, reinforcing the potential value of pathway-centered analyses and AI-assisted genomic interrogation for identifying subtype-specific biological signatures in CTCL.

## 4. Discussion

CTCL encompasses a biologically heterogeneous group of malignancies characterized by clonal expansion of skin-homing T lymphocytes with diverse clinical behaviors and molecular architectures. Among CTCL subtypes, SS represents a particularly aggressive leukemic variant distinguished by erythroderma, circulating malignant T cells, and poor clinical outcomes. Although prior genomic studies have identified recurrent mutations affecting T-cell signaling, epigenetic regulation, and immune pathways in CTCL, the extent to which pathway-level differences distinguish SS from other CTCL subtypes has remained incompletely characterized. In this study, we used a pathway-centric analytical framework augmented by conversational artificial intelligence (AI) to systematically compare the somatic mutational landscape of SS and non-SS CTCL. Our results demonstrate that SS is not defined by a higher overall mutational burden but rather by qualitative differences in the biological pathways affected, particularly those governing epigenetic regulation, transcriptional control, immune signaling, and apoptosis.

One of the most prominent findings in our analysis was the enrichment of mutations affecting epigenetic regulators in SS, including genes such as TET2, CREBBP, CHD3, and BRD9. Epigenetic dysregulation is increasingly recognized as a hallmark of T-cell lymphomagenesis, influencing chromatin accessibility, transcriptional programs, and lineage-specific gene expression. Alterations in epigenetic modifiers may contribute to the aberrant transcriptional states observed in malignant T cells, enabling immune evasion and sustained proliferation. Previous studies have reported recurrent mutations in chromatin-remodeling genes in CTCL, suggesting that disruption of epigenetic regulatory networks may play a central role in disease progression. Our findings reinforce this concept and suggest that epigenetic remodeling may be particularly important in the pathogenesis of SS, potentially contributing to its systemic and leukemic phenotype.

In addition to epigenetic regulators, SS tumors showed relatively greater involvement of tumor suppressor and cell-cycle regulatory pathways, including alterations affecting TP53 and related checkpoint genes. Loss of tumor suppressor function is a common driver of genomic instability and malignant progression in many hematologic malignancies. In CTCL, disruption of these regulatory circuits may facilitate clonal expansion of malignant T cells and enable escape from normal apoptotic controls. Notably, the coexistence of tumor suppressor loss and epigenetic dysregulation in SS suggests a model in which transcriptional reprogramming and impaired checkpoint control cooperate to promote aggressive disease behavior.

Another pathway category enriched in SS involved NFAT signaling and proximal T-cell receptor (TCR) activation pathways, including mutations in genes such as PLCG1 and PRKG1. NFAT transcription factors play a critical role in T-cell activation, differentiation, and cytokine production. Dysregulation of NFAT signaling has been implicated in several T-cell malignancies and may contribute to aberrant activation states in malignant T cells. Mutations affecting TCR signaling components can lead to constitutive downstream signaling, promoting proliferation and survival of malignant clones. The enrichment of NFAT-related alterations in SS suggests that aberrant T-cell activation programs may represent a key biological feature of the disease, potentially contributing to the widespread immune dysregulation observed clinically in these patients.

Our analysis also identified alterations affecting genes involved in apoptosis and immune regulation, including PDCD1 and FAS-associated pathways, in SS tumors. These genes regulate immune checkpoint signaling and programmed cell death pathways that normally maintain immune homeostasis. Disruption of these regulatory mechanisms may enable malignant T cells to evade immune surveillance while resisting apoptosis. The observation that apoptosis and immune regulatory genes were mutated exclusively in the SS cohort, although at low frequency, suggests that immune escape mechanisms may contribute to the distinct clinical behavior of SS, which is characterized by systemic dissemination and profound immune dysregulation.

In contrast, non-SS CTCL subtypes demonstrated relatively greater involvement of MAPK and JAK-STAT signaling pathways, including mutations affecting MAPK1, BRAF, STAT3, and JAK3. Both signaling cascades play central roles in cytokine signaling, cellular proliferation, and survival in lymphoid cells. Activation of the MAPK pathway has been implicated in several T-cell lymphomas, and mutations in RAS-RAF-MEK signaling components may sensitize tumors to targeted inhibition of the MAPK cascade. Similarly, dysregulation of JAK-STAT signaling has been widely reported in CTCL and other T-cell malignancies, reflecting the importance of cytokine-mediated signaling in T-cell biology. The relative enrichment of these pathways in non-SS CTCL suggests that different CTCL subtypes may rely on distinct oncogenic signaling programs, with MAPK and JAK-STAT pathways potentially representing more prominent drivers outside the SS phenotype.

Beyond individual gene and pathway alterations, our co-mutation analysis revealed distinct patterns of genomic interaction across CTCL subtypes. The SS cohort exhibited a comparatively restricted network of gene-gene interactions, suggesting that a smaller set of cooperating alterations may define the molecular architecture of the disease. In contrast, non-SS CTCL demonstrated a broader co-mutation network involving numerous gene pairs. These observations imply that SS may evolve through a more constrained set of transcriptional and immune-regulatory dependencies, whereas non-SS CTCL may arise through more heterogeneous mutational combinations spanning multiple signaling pathways. This difference in network topology may reflect distinct evolutionary pressures shaping tumor development in each subtype.

Importantly, despite these pathway-level differences, we observed no significant difference in tumor mutational burden (TMB) between SS and non-SS CTCL. This finding indicates that the biological distinction between these disease subtypes is not simply attributable to increased genomic instability or mutation accumulation. Instead, our results suggest that the identity and functional context of somatic mutations are more critical than their absolute number in determining CTCL subtype biology. This concept aligns with emerging views in cancer genomics emphasizing pathway-level dysregulation rather than mutation quantity as a key determinant of tumor phenotype.

Several genes showed borderline subtype-specific enrichment, including ERBB2, WWC1, POSTN, GUCY2F, and PCDHB13, which were observed exclusively in the non-SS cohort. Although these associations did not reach conventional statistical significance, their consistent presence in non-SS CTCL raises the possibility that they may contribute to subtype-specific biology. For example, ERBB2 encodes a receptor tyrosine kinase involved in growth factor signaling and oncogenic transformation in multiple tumor types. Similarly, POSTN encodes periostin, a secreted extracellular matrix protein implicated in tumor microenvironment remodeling and metastatic progression. Future studies using larger CTCL cohorts will be necessary to determine whether these genes represent true subtype-specific drivers or biomarkers.

A unique aspect of this study was the integration of conversational AI-assisted analysis within the genomic discovery workflow. The AI platform facilitated rapid exploration of mutation datasets, prioritization of biologically coherent pathway patterns, and interactive interpretation of genomic relationships. Importantly, this approach did not replace conventional statistical analysis but served as a complementary tool to accelerate hypothesis generation and data interpretation. In the context of rare malignancies such as CTCL, where datasets are often small and heterogeneous, AI-assisted analytical frameworks may provide valuable support for identifying biologically meaningful signals that warrant further investigation.

This study has several limitations. First, the analysis was conducted using a relatively small cohort derived from publicly available genomic data, which limits statistical power for detecting subtype-specific associations. Second, the cross-sectional design of the dataset prevents direct inference regarding the temporal evolution of mutational events during disease progression. Third, the non-SS cohort included multiple CTCL subtypes with distinct clinical and biological characteristics, which may contribute to additional heterogeneity. Finally, although pathway-level annotation provides valuable biological context, functional validation experiments are required to confirm the mechanistic significance of these genomic alterations.

Despite these limitations, our findings provide several important insights into the molecular biology of CTCL. By applying a pathway-centric analytical framework, we demonstrate that SS is characterized by disproportionate involvement of epigenetic, transcriptional, and immune-regulatory pathways, whereas non-SS CTCL subtypes show relatively greater activation of MAPK and JAK-STAT signaling networks. These results highlight the importance of examining genomic alterations within their broader biological pathways rather than focusing solely on individual genes.

In summary, our study provides a systematic comparison of pathway-level mutational patterns between SS syndrome and other CTCL subtypes. The results suggest that the aggressive clinical phenotype of SS may arise from cooperative dysregulation of epigenetic programs, tumor suppressor pathways, T-cell signaling, and immune regulatory mechanisms, rather than from increased mutational burden. These findings generate testable hypotheses for future functional studies and underscore the potential of AI-assisted analytical frameworks to accelerate discovery in translational cancer genomics.

## Data Availability

All data used in the present study is publicly available at https://www.cbioportal.org/. The datasets used in our study were aggregated/summary data, and no individual-level data were used. Additional data can be provided upon reasonable request to the authors.

---

**Table S1.** Gene-level comparison of borderline-significant somatic mutation frequencies between Sézary syndrome and non-Sézary cutaneous T-cell lymphoma cohorts.

| Gene | Sézary Syndrome Mutated n (%) | Sézary Syndrome Wild-Type n (%) | Non-Sézary Syndrome Mutated n (%) | Non-Sézary Syndrome Wild-Type n (%) | p-value |
| --- | --- | --- | --- | --- | --- |
| ERBB2 | 0 (0.0%) | 26 (100.0%) | 3 (17.6%) | 14 (82.4%) | 0.0551 |
| GUCY2F | 0 (0.0%) | 26 (100.0%) | 3 (17.6%) | 14 (82.4%) | 0.0551 |
| WWC1 | 0 (0.0%) | 26 (100.0%) | 3 (17.6%) | 14 (82.4%) | 0.0551 |
| PCDHB13 | 0 (0.0%) | 26 (100.0%) | 3 (17.6%) | 14 (82.4%) | 0.0551 |
| POSTN | 0 (0.0%) | 26 (100.0%) | 3 (17.6%) | 14 (82.4%) | 0.0551 |

**Figure S1.**
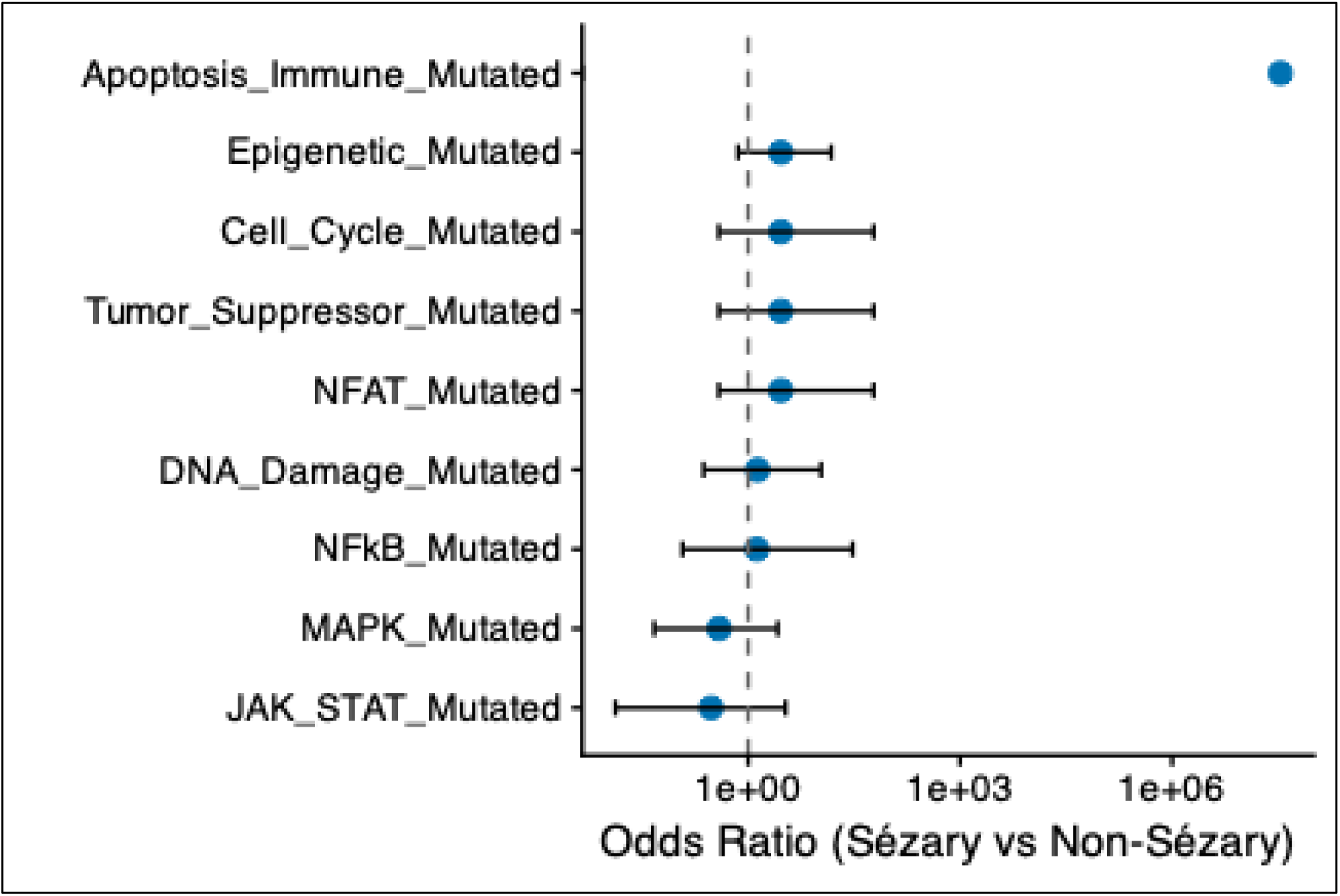
Forest plot of pathway-level mutation enrichment comparing Sézary syndrome and non-Sézary cutaneous T-cell lymphoma. This forest plot summarizes odds ratios and corresponding confidence intervals for pathway-level mutation enrichment between Sézary syndrome (SS) and non-Sézary syndrome (non-SS) cutaneous T-cell lymphoma (CTCL) tumors. Odds ratios greater than one indicate enrichment in SS, whereas values below one indicate enrichment in non-SS CTCL. Alterations in MAPK and JAK/STAT signaling pathways show odds ratios modestly below unity, suggesting relatively higher mutation frequencies in non-SS tumors. In contrast, mutations affecting epigenetic regulators, cell-cycle control genes, tumor suppressor pathways, and NFAT signaling demonstrate odds ratios greater than one, indicating increased prevalence in Sézary syndrome. DNA damage response and NF-κB pathway alterations display odds ratios close to unity, consistent with comparable mutation frequencies across CTCL subtypes. Notably, mutations in apoptosis and immune regulatory genes exhibit the strongest association with SS, reflected by a markedly elevated odds ratio resulting from the absence of such mutations in the non-SS cohort. Overall, these findings suggest that while several signaling pathways are shared between CTCL subtypes, SS shows a distinct enrichment of alterations involving epigenetic regulation, cell-cycle control, tumor suppressor pathways, NFAT signaling, and immune regulatory mechanisms.

**Figure S2.**
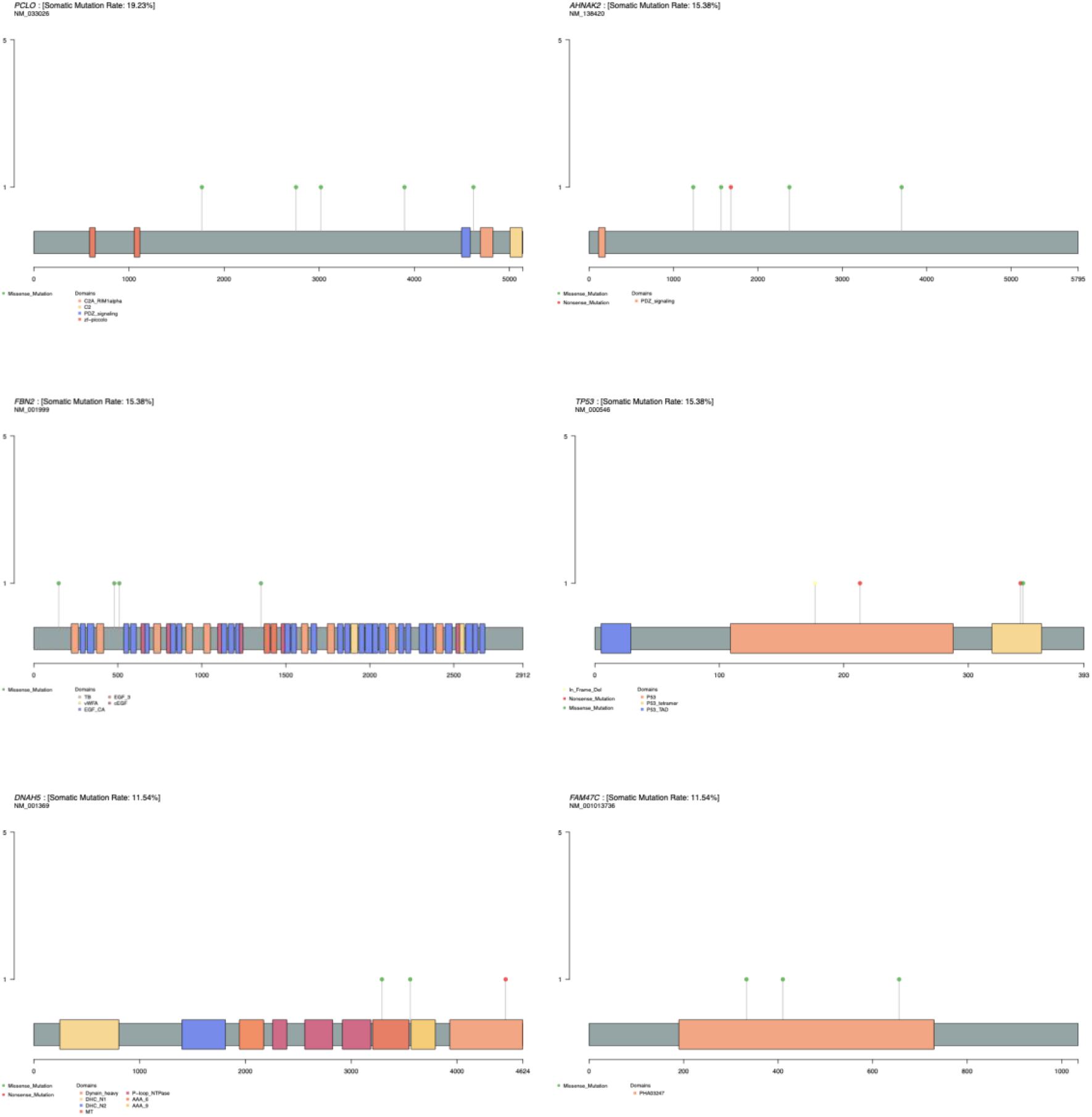
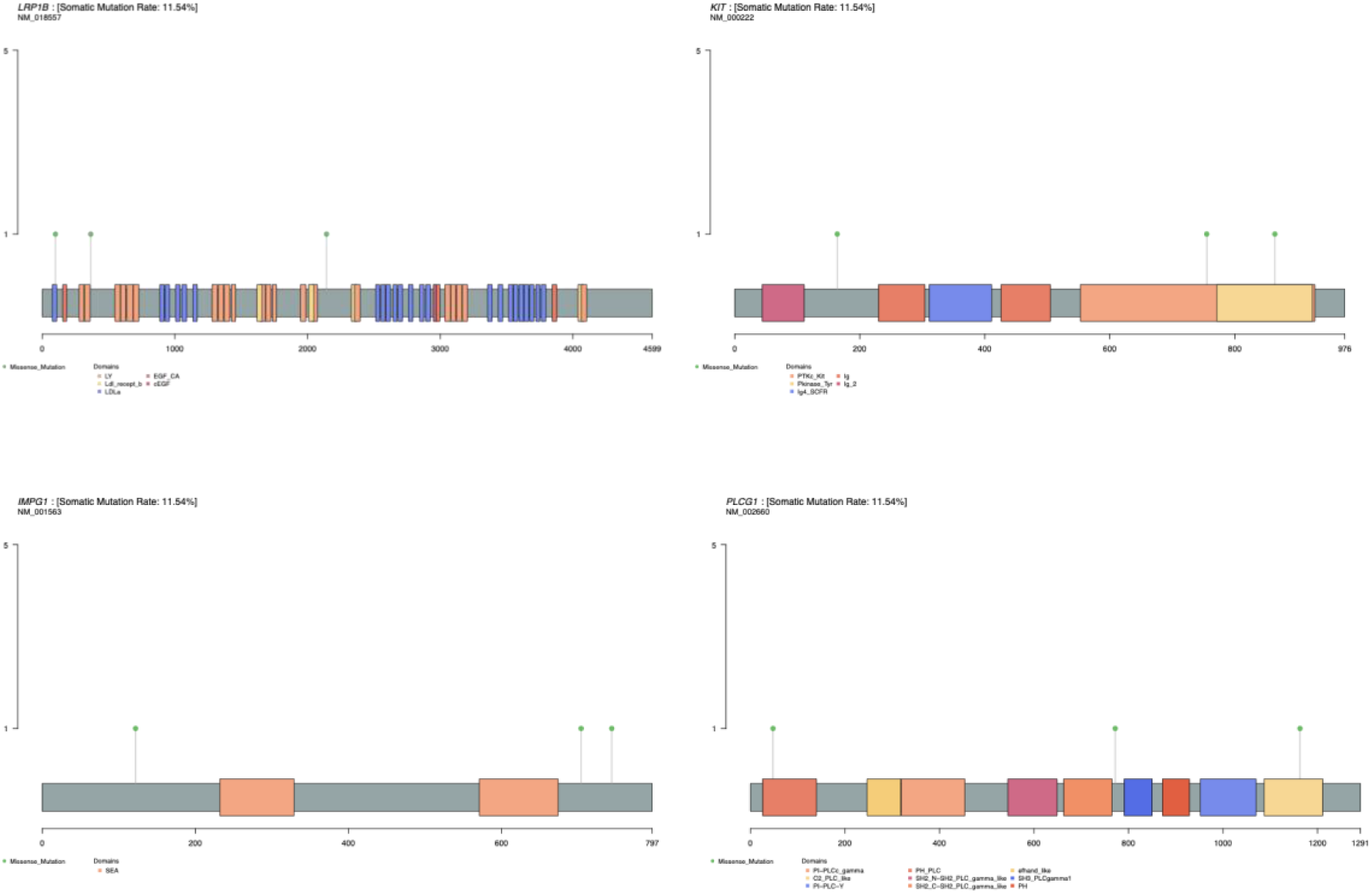
Lollipop plots illustrating the distribution of somatic mutations in the top 10 recurrently mutated genes in the Sézary syndrome cohort. This figure presents lollipop plots depicting the positional distribution of somatic mutations across the protein sequences of the ten most frequently mutated genes identified in the Sézary syndrome (SS) cohort. Each panel corresponds to a single gene, and each lollipop marker represents an individual mutation event. The horizontal axis denotes the amino acid position along the protein sequence, while the vertical lollipop height reflects the number of tumor samples harboring mutations at that site. Colored markers indicate the type of somatic alteration, and annotated protein domains are shown along the protein structure to contextualize mutation locations. Together, these plots highlight mutation clustering and potential hotspot regions across the most frequently altered genes in SS, providing insight into candidate functional domains that may contribute to disease pathogenesis.

**Figure S3.**
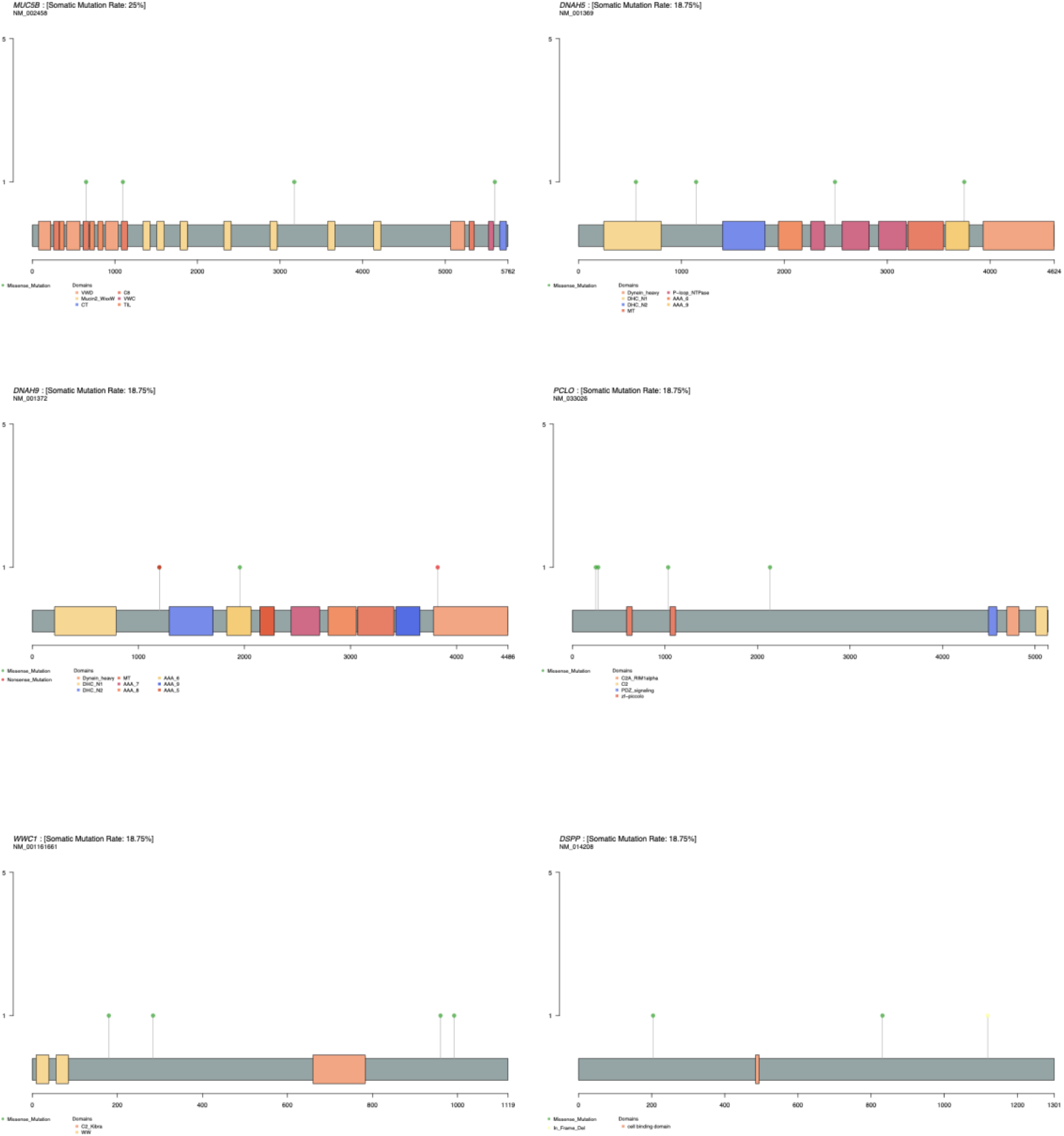
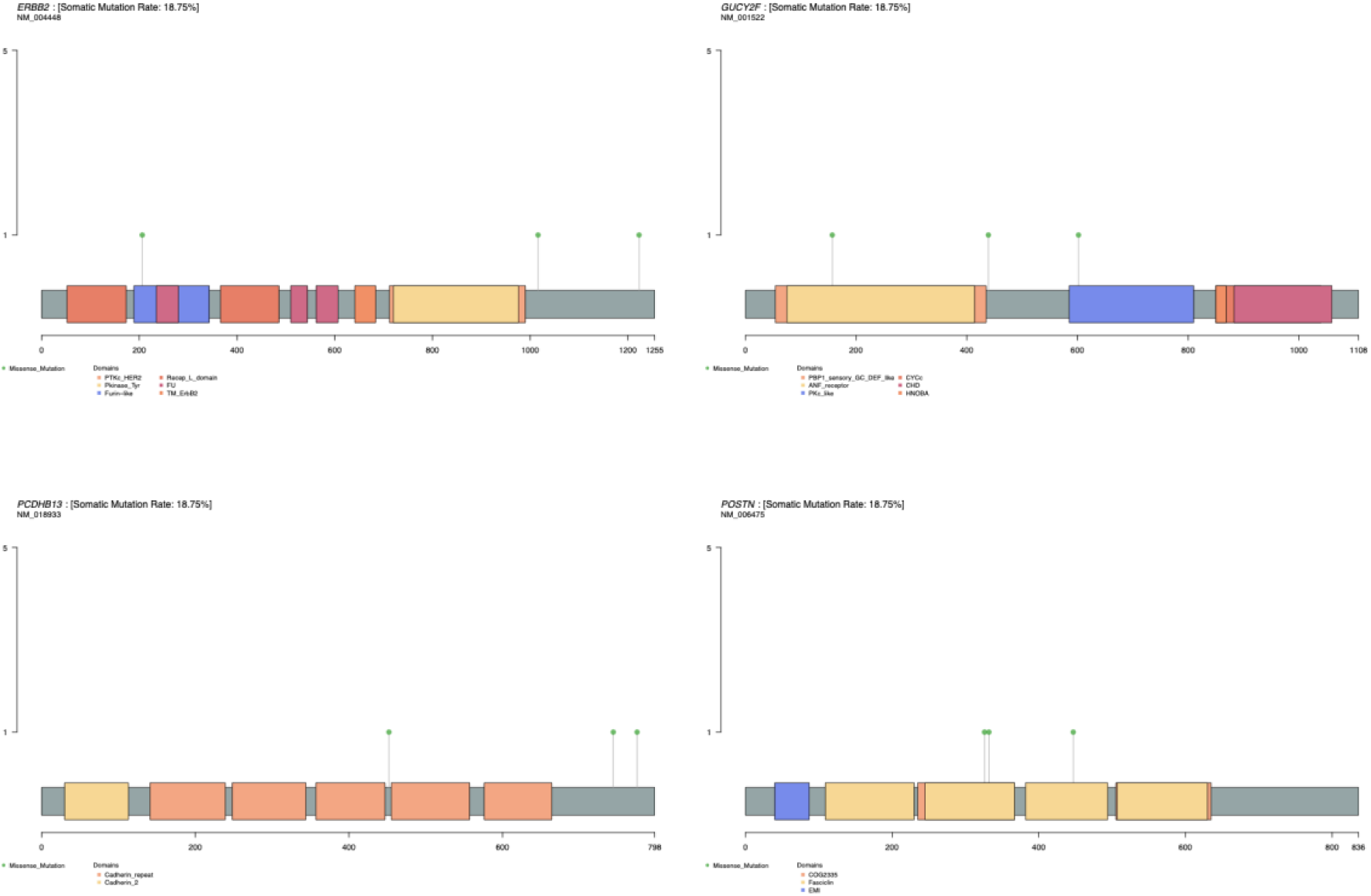
Lollipop plots illustrating the distribution of somatic mutations in the top 10 recurrently mutated genes in the non-Sézary cutaneous T-cell lymphoma cohort. This figure presents lollipop plots depicting the positional distribution of somatic mutations across the protein sequences of the ten most frequently mutated genes identified in the non-Sézary (non-SS) cutaneous T-cell lymphoma (CTCL) cohort. Each panel corresponds to an individual gene, with lollipop markers representing mutation events observed across tumor samples. The horizontal axis indicates the amino acid position along the protein sequence, while the vertical height of each lollipop reflects the number of samples harboring mutations at that specific site. Colored markers denote different mutation types, and annotated protein domains provide structural context for the observed alterations. These visualizations highlight mutation distribution patterns and potential hotspot regions across recurrently altered genes in non-SS CTCL, offering insight into candidate functional regions that may contribute to disease biology.

**Figure S4.**
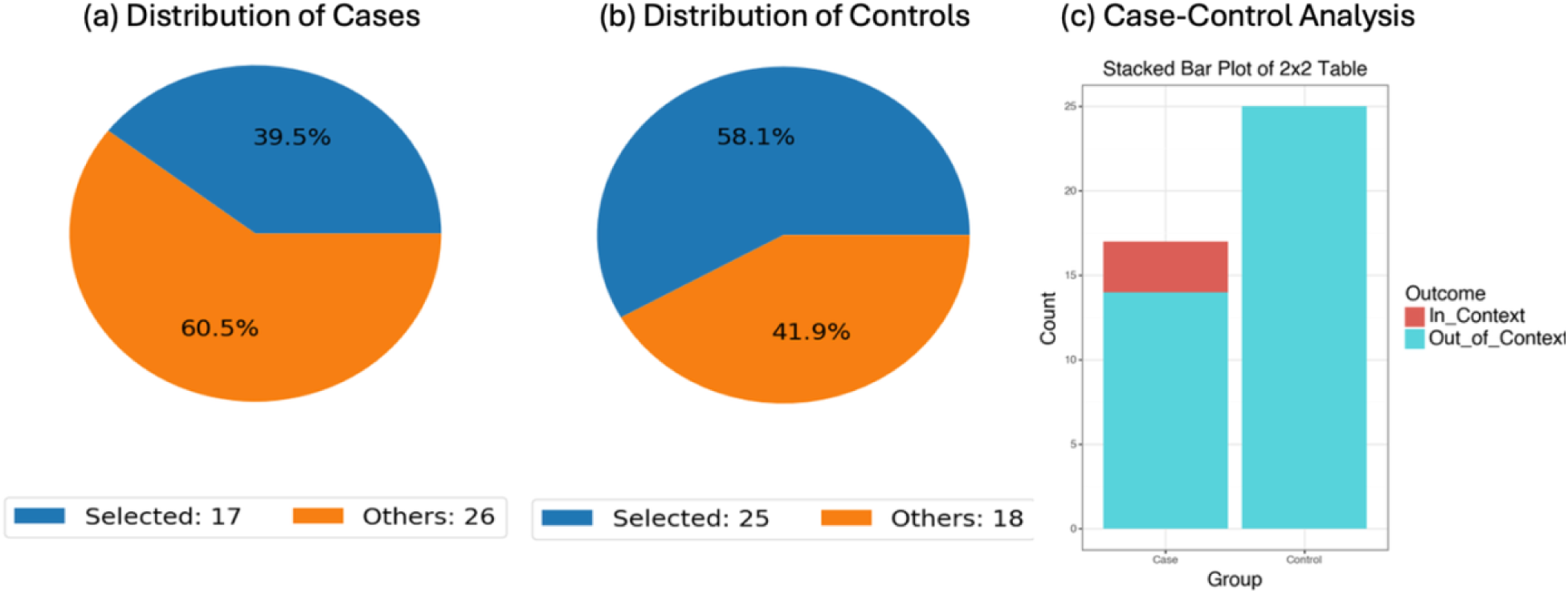
Conversational AI-assisted case-control analysis of ERBB2 mutation prevalence in Sézary syndrome versus non-Sézary cutaneous T-cell lymphoma. This figure illustrates a conversational AI-guided case-control framework used to evaluate whether ERBB2 mutations are differentially represented between CTCL subtypes. The analysis compares a case cohort of non-Sézary (non-SS) cutaneous T-cell lymphoma (CTCL) samples (n = 17) with a control cohort of Sézary syndrome (SS) samples (n = 25) derived from the Columbia University CTCL dataset available through cBioPortal. Cohorts were defined using structured query logic within the conversational AI analytical environment. Panels (a) and (b) display pie charts summarizing the distribution of selected samples within the broader dataset for the case and control cohorts, respectively. Panel (c) presents a stacked bar plot summarizing the number of ERBB2-mutated (“in-context”) and non-mutated (“out-of-context”) samples in each group based on a 2×2 contingency framework. ERBB2 mutations were observed in 3 of 17 non-SS CTCL cases and 0 of 25 SS samples. Fisher’s exact test was used to assess statistical significance of the association between ERBB2 mutation status and disease subtype. The odds ratio estimate suggested a potential enrichment of ERBB2 alterations in non-SS CTCL, although the confidence interval was wide due to the absence of ERBB2 mutations in the SS cohort. These results illustrate how conversational AI-guided analytical workflows can rapidly generate case-control comparisons of genomic alterations, facilitating exploratory hypothesis testing and visualization of mutation frequency differences across CTCL subtypes.

**Figure S5.**
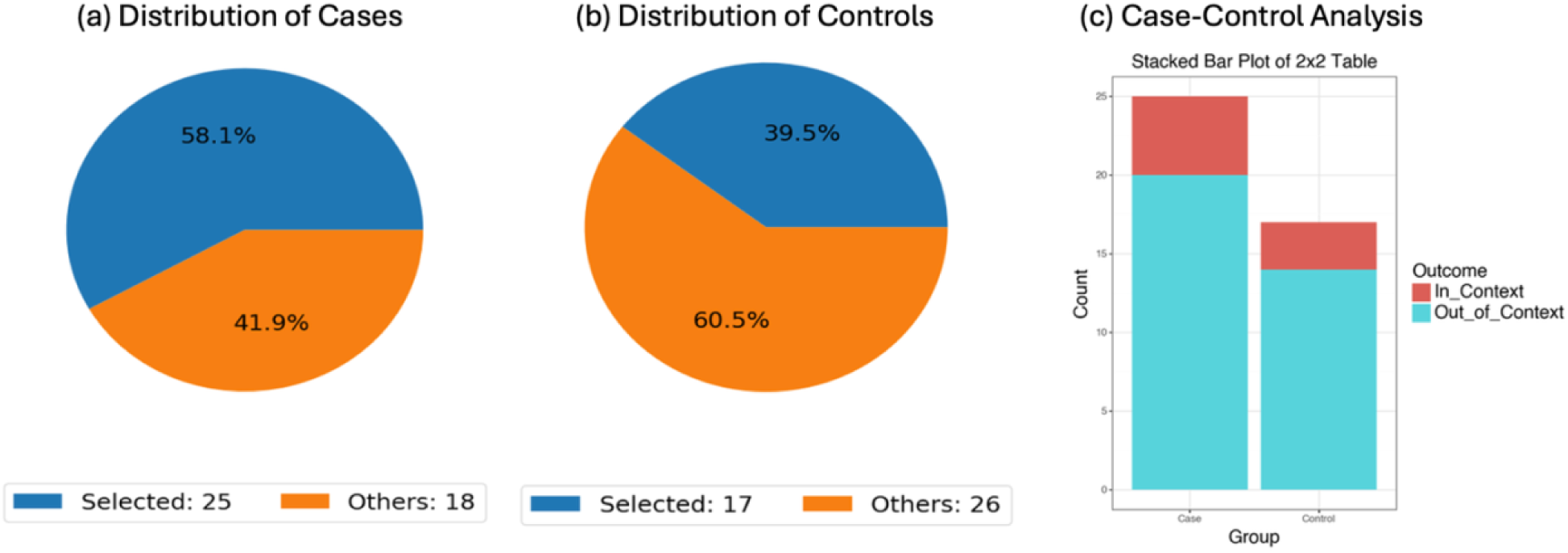
Conversational AI-guided case-control evaluation of PCLO mutation prevalence in Sézary syndrome versus non-Sézary cutaneous T-cell lymphoma. This figure presents a conversational AI-driven case-control analysis assessing whether PCLO mutations differ in prevalence between Sézary syndrome (SS) and non-Sézary (non-SS) CTCL tumors. Using structured cohort definitions within the AI analytical framework, the case cohort consisted of SS samples (n = 25) and the control cohort included non-SS CTCL samples (n = 17) derived from the Columbia University CTCL dataset available through cBioPortal. Panels (a) and (b) illustrate the relative distribution of selected samples within the case and control cohorts compared with the total dataset. Panel (c) displays a stacked bar plot summarizing PCLO-mutated (“in-context”) and non-mutated (“out-of-context”) samples using a 2×2 contingency table. PCLO mutations were detected in 5 of 25 SS samples (20.0%) and 3 of 17 non-SS CTCL samples (17.65%). Statistical comparison using Fisher’s exact test indicated no significant difference in PCLO mutation frequency between the two groups (p = 1.0). The estimated odds ratio was 1.17, with a wide confidence interval reflecting the limited sample size. These results suggest that PCLO mutation prevalence is comparable between SS and non-SS CTCL within this dataset. The analysis highlights the ability of conversational AI-enabled analytical workflows to rapidly construct case-control genomic comparisons and visualize mutation frequency patterns across CTCL subtypes.

**Figure S6.**
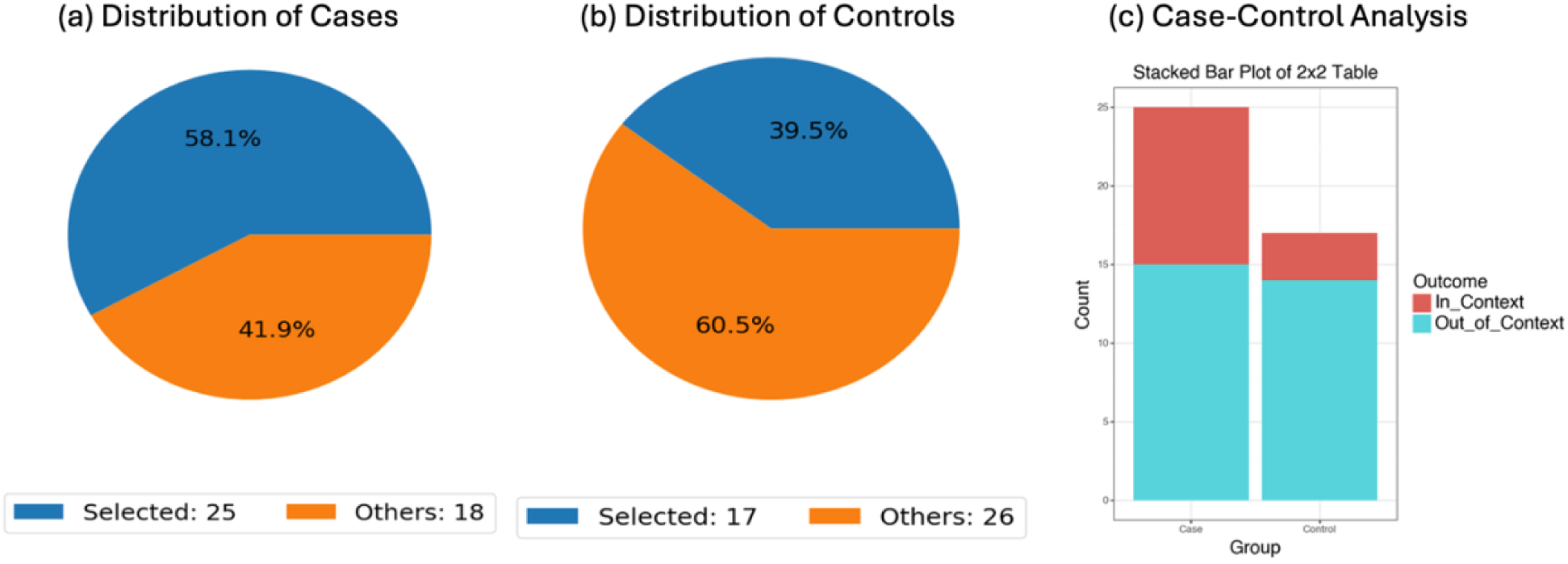
Conversational AI-enabled case-control comparison of epigenetic pathway mutations in Sézary syndrome and non-Sézary cutaneous T-cell lymphoma. This figure depicts an AI-assisted case-control analysis examining whether mutations in epigenetic regulatory genes are differentially represented between Sézary syndrome (SS) and non-Sézary syndrome (non-SS) cutaneous T-cell lymphoma (CTCL) tumors. Cohorts were defined using structured criteria within the conversational AI analytical workflow, with the case cohort consisting of SS samples (n = 25) and the control cohort including non-SS CTCL samples (n = 17) derived from the Columbia University CTCL dataset available through cBioPortal. Panels (a) and (b) display pie charts illustrating the proportion of selected samples relative to the total dataset within the case and control cohorts. Panel (c) presents a stacked bar plot summarizing the number of tumors harboring epigenetic gene alterations (“in-context”) versus those lacking such alterations (“out-of-context”) across both groups using a 2×2 contingency framework. Epigenetic pathway mutations were identified in 10 of 25 SS syndrome samples (40.0%) and 3 of 17 non-SS CTCL samples (17.65%). Statistical comparison using Fisher’s exact test yielded no statistically significant difference between cohorts (p = 0.231), although the calculated odds ratio of 3.11 suggests a trend toward enrichment of epigenetic alterations in SS. These results highlight the application of conversational AI-driven analytical pipelines for rapidly constructing genomic case-control comparisons and visualizing pathway-level mutation patterns across CTCL subtypes.

